# Multiomics reveals gut dysbiosis links to fatty acid dysmetabolism in early phase of acute myocardial infarction

**DOI:** 10.1101/2024.05.21.24307709

**Authors:** Jiajun Ying, Yong Fan, Ning Huangfu, Kewan He, Teng Hu, Pengpeng Su, Xintao Hu, Hequn He, Wei Liang, Junsong Liu, Jinsong Cheng, Shiqi Wang, Ruochi Zhao, Hengyi Mao, Fuwei He, Jia Su, Honglin Zhou, Zhenwei Li, Xiaohong Fei, Xiafei Sun, Peipei Wang, Minfang Guan, Weiping Du, Shaoyi Lin, Yong Wang, Fangkun Yang, Renyuan Fang, Ziqing Kong, Xiaomin Chen, Hanbin Cui, Jiajun Ying and Yong Fan contributed equally to this work

**Author notes:** Corresponding author: X.C. or H.C.

## Abstract

**Background:** Acute myocardial infarction (AMI) remains a major cause of death, with limited understanding of its early risk stratification. While gut microbiome disturbances has been associated with late-stage AMI, the connection to early-stage AMI (eAMI) is less explored.

**Methods:** Using metabolomics and metagenomics, we analyzed 56 samples, comprising 30 eAMI patients (within 12 hours of onset) and 26 age- and gender-matched healthy controls, to discern the influence of gut microbes and their metabolites.

**Results:** We found the eAMI plasma is dominated by increased long-chain fatty acids (LCFAs), 14 of which provide differentiating power of eAMI patients from HCs. Multiomics analysis reveals up to 70% of the variance in LCFAs of eAMI patients can be explained by altered gut microbiome. Higher-resolution profiling of gut bacterial species demonstrated that bacterial structural variations are mechanistically linked to LCFAs dysregulation. By *in silico* molecular docking and *in vitro* thrombogenic assay in isolated human platelets, we highlighted that eAMI-associated LCFAs contribute to platelet aggregation, a driving factor for AMI initiation.

**Conclusions:** LCFAs hold significant potential as early biomarkers of AMI and gut microbiome contributes to altered LCFAs in eAMI. Further studies are imperative to expand upon these observations to better leverage LCFAs as a potential biomarker for eAMI and as a therapeutic target for inhibition of platelet aggregation in eAMI.

## Introduction

Acute myocardial infarction (AMI), the most lethal type in coronary artery disease (CAD) and primary cause leading to human mortality and morbidity(1–3), is mainly resulted from the rupture or erosion of atherosclerotic plaque, accompanied by the formation of an occlusive or sub-occlusive thrombus in one or more coronary artery branches(4,5). Accordingly, AMI is divided into two subtypes, ST-segment elevation myocardial infarction (STEMI) and non-ST-segment elevation myocardial infarction (NSTEMI). It has been showed that systemic or local inflammation, imbalance in adaptive immune pathways can modulate atherosclerotic plaque activity and contribute to coronary dysfunction and myocardial ischemic necrosis(5–7). However, the exact underlying mechanism responsible for the progression of coronary artery disease and occurrence of adverse event remains unclear.

A rising body of epidemiological study suggests that gut microbiota have a pivotal role in low-grade inflammatory metabolic diseases such as obesity(8) and diabetes(9), which are considered risk factor for atherosclerotic development. Thereafter, pathological evidence indicates that bacteria from the gut or the oral cavity might translocate to and reside in atherosclerotic plaques and could affect the development of atherosclerotic cardiovascular disease (ASCVD)(10,11). Improved sequencing technology and bioinformatics analyses have provided the several significant associations between the microbiota taxa and ASCVD(12,13).

On how gut microbiota promote to the development of ASCVD, we have summarized recent advances in the atherosclerosis-linked alterations in gut microbiome, highlighting the microbiota-related circulating metabolites that accelerate the development of ASCVD(14). Microbiota-synthesized metabolites such as trimethylamine N-oxide, secondary bile acids, short-chain fatty acids, lipopolysaccharides and phenylacetylglutamine have been recognized as the important contributor to atherosclerotic consequences. Nevertheless, whether gut microbiota participate in the initiation of AMI, some observational study reported the statistical correlation between gut bacteria profiling and AMI clinical phenotype. However, results are inconsistent across previous findings(15,16).

So far, the reported investigations on the associations between intestinal microbes and AMI recruited mostly patients in the late or stable stage of AMI, sparse report on very early phase of disease has been shown. Whether gut microbes contribute to the early stage of AMI (eAMI) lesion through its derived metabolites remains unknown. Here, we recruited a cohort of 30 patients with eAMI within 12 hours and 26 normal controls, for whom we performed metagenomic sequencing of the gut microbiome and plasma metabolomics. We found that long-chain fatty acids (LCFAs) dominate the metabolome perturbations in eAMI plasma, and the gut microbiome at species level explains up to 70% of the variance in LCFAs composition in eAMI patients. We also found that eAMI-associated LCFAs may possess potent thrombogenic functionality by molecular docking and platelet aggregation assay, which furtherly supports the rational for causal relationship between altered gut microbiota and the initiation of AMI.

## Materials and methods

### Study design and cohort description

This study enrolled 30 AMI patients and 26 normal controls who were hospitalized in the First Affiliated Hospital of Ningbo University from February 2021 to November 2021. Participants were in strictly conformity with the inclusion and exclusion criteria (**Figure S1**). The diagnosis of AMI is based on the guidelines of the American Heart Association (ACC) in 2021(17).

Inclusion criteria were diagnoses of AMI defined as follows: cardiac troponin (cTn) I/T > the upper limit of the 99% normal reference value and have any one or more of the following: significant chest pain of myocardial ischemia; changes in newly diagnosed ischemic electrocardiogram (ST-T changes or left bundle branch block or pathological Q wave); imaging evidence suggested new loss of myocardial activity or regional wall motion abnormalities; abnormal coronary angiography.

Exclusion criteria were: history of digestive diseases; severe hepatic and renal dysfunction; history of autoimmune diseases, malignant hematological diseases and tumor; infection within one month of the study or the use of antibiotic or probiotic. Normal controls were hospitalized for health examination at the same period.

The study protocol was approved by the Ethics Committee of The First Affiliated Hospital of Ningbo University (No. 2021-R018-YJ01). The study was performed in accordance with the principles of the Declaration of Helsinki. All participants provided written, informed consent for participation in the study.

### Sample collection and biochemical analysis

Peripheral venous blood was drawn immediately after the patients arrived at the emergency department, or first moments after hospitalization for normal control individuals. Blood samples were centrifuged immediately at 4 °C, 4000 rpm for 10 minutes, and the supernatant (plasma) was stored at −80 °C for further analysis. The first morning fresh fecal (each 400-600 mg) and urine samples were collected from all the participants under the hospital diet, then placed into sterile cryotube and frozen at −80°C refrigerator before further use.

Whole blood samples for biochemical measurements were collected after at least 10-hour fasting period. The measurements of fasting glucose, lactate dehydrogenase, creatine kinase, c-reactive protein, serum alanine aminotransferase, aspartate aminotransferase, glutathione S-transferase, creatinine, glycated serum protein, high-density lipoprotein cholesterol, low-density lipoprotein cholesterol, apolipoproteins, lipoprotein, and serum amyloid A were performed by an autoanalyzer (AU5800, Beckmancoulter). HbA1C was measured by an automated clinical analyzer (H100plus, Lifotronic). Plasma brain natriuretic peptide level was measured by a full-automatic chemiluminescence immunoassay analyzer (Cobas e602, Roche). Plasma troponin was measured using an autoanalyzer (DXI800, Beckmancoulter). Urine samples were analyzed by urine sediment analyzer (UF-5000, Sysmex).

### Pseudo-targeted metabolomics profiling and preprocessing

Metabolomic profiling was performed utilizing the CalOmics metabolomics platform at Calibra Lab, affiliated with DI^’^AN Diagnostics. The analytical instrumentation comprised an ACQUITY 2D UPLC system (Waters, Milford, MA, USA) coupled with a Q Exactive (QE) hybrid Quadrupole-Orbitrap mass spectrometer (Thermo Fisher Scientific, San Jose, USA). Metabolites were analyzed, based on the chemical properties of the serum metabolites, utilizing both polar ionic and lipid modes.

Sample preparation involved the extraction of metabolites using a methanol solvent at a 1:4 ratio. Following extraction, mixtures were shaked for three minutes and subsequently subjected to centrifugation at 4000 × g for 10 minutes at 20°C to precipitate proteins. The resulting 100 μL aliquots of supernatant were transferred onto sample plates, dried under a stream of nitrogen, and reconstituted in appropriate solvents for injection into the UPLC-MS/MS system.

The QE mass spectrometer settings included a mass resolution of 35,000 and a scan range from 70 to 1000 m/z. For the first UPLC-MS/MS method, the instrument operated in positive electrospray ionization (ESI) mode using a C18 reverse-phase UPLC column (UPLC BEH C18, 2.1×100 mm, 1.7 μm; Waters). The mobile phases consisted of water (A) and methanol (B) with 6.5 mM ammonium bicarbonate at pH 8 for gradient elution. The third method similarly employed the C18 reverse-phase column in ESI positive mode, but used a mobile phase of water (A) and a mixture of methanol, acetonitrile, and water (B) containing 0.05% perfluoropentanoic acid (PFPA) and 0.01% formic acid (FA). In the fourth method, the system was operated in negative ESI mode with a hydrophilic interaction liquid chromatography (HILIC) column (UPLC BEH Amide, 2.1×150 mm, 1.7 μm; Waters), and the mobile phases were water (A) and acetonitrile (B) containing 10 mM ammonium formate.

Following the preprocessing of raw data and rigorous data quality control assessments, ion peaks were extracted employing proprietary in-house information technology hardware and software systems. Metabolites were subsequently identified through querying a bespoke library, which was compiled using pure standards sourced from commercial providers or other available channels. The identification protocol for metabolites within the samples was stringent, requiring a precise match across three specific criteria when compared to the library entries: a narrow retention index window, mass accuracy within a 10 ppm deviation, and tandem mass spectrometry (MS/MS) spectra that exhibit high scores in both forward and reverse searches. Quantification of each metabolite was achieved by calculating the peak area using the area-under-the-curve (AUC) method. This approach ensures accurate measurement of metabolite concentrations within the samples.

Before statistical analysis, raw peak areas were normalized to adjust for system fluctuation among different run days. The normalized peak areas were then log-transformed (log2) to reduce data distribution skewness and be in approximate normal distribution (Gaussian distribution). Missing values in peak area matrix were imputed by using the minimal detection value of a metabolite among all tested samples.

### DNA extraction and library construction of fecal samples

Bacterial DNA was extracted from fecal samples using the Qiagen DNeasy PowerSoil Kit (Cat. No.: 47014) according to the manufacturer’s protocol. Stool samples were lysed through a combination of chemical and mechanical homogenization. Lysis buffer was added to a zirconium bead tube containing the sample, followed by bead beating, which was performed using a standard benchtop vortex with a bead tube adapter. The resulting crude lysate underwent inhibitor removal to ensure sample cleanup. Subsequently, the purified lysate was mixed with an equal volume of DNA binding solution and passed through a silica spin filter membrane. The membrane was then subjected to a two-step washing process. Finally, the silica-bound DNA was eluted using a 10 mM Tris elution buffer.

For polymerase chain reaction (PCR) analysis, the samples were processed at LC-BIO Technologies Co., Hangzhou, China. The construction of the DNA library was accomplished using the TruSeq Nano DNA LT Library Preparation Kit (FC-121-4001). Fragmentation of the DNA was performed using dsDNA Fragmentase (NEB, M0348S) at 37°C for 30 minutes. This step produced blunt-ended DNA fragments via a combination of fill-in reactions and exonuclease activity. Size selection was then conducted using the sample purification beads provided in the kit.

To prepare the DNA fragments for adapter ligation, an adenine (A) base was added to the blunt ends of each strand. Each adapter, designed with a thymine (T) base overhang, facilitates the ligation to the A-tailed DNA fragments. These adapters are crucial as they contain necessary sites for the hybridization of sequencing primers, enabling single, pair-end, and indexed sequencing reads.

Ligation involved either single or dual index adapters, and the products were subsequently amplified via PCR. The amplification parameters included an initial denaturation at 95°C for 3 minutes, followed by 8 cycles of denaturation at 98°C for 15 seconds, annealing at 60°C for 15 seconds, extension at 72°C for 30 seconds, and a final extension at 72°C for 5 minutes. This process prepares the library for high-throughput sequencing analyses.

### Metagenomic sequencing data processing

Raw sequencing data were processed to generate clean reads suitable for downstream analyses. Initially, sequencing adapters were excised from the reads using Cutadapt (version 1.9). Subsequently, low-quality reads were trimmed using fqtrim (version 0.94), which employs a sliding-window algorithm. Reads were then aligned to the host genome using Bowtie2 (version 2.2.0) to eliminate host-derived sequences. Following the quality filtering, the remaining reads were assembled de novo using IDBA-UD (version 1.1.1) to construct individual metagenomes for each sample. Predictions of coding regions (CDS) within the metagenomic contigs were performed by MetaGeneMark (version 3.26). The CDS sequences across all samples were clustered using CD-HIT (version 4.6.1) to generate a set of unigenes. Unigene abundance in each sample was estimated using transcripts per million (TPM) based on read alignment performed again with Bowtie2 (version 2.2.0). Taxonomic classification of the unigenes was achieved by aligning them to the NCBI non-redundant (NR) database using DIAMOND (version 0.9.14). The taxonomic annotations and abundance profiles of these unigenes were utilized to conduct differential analysis at both taxonomic and gene-specific levels. Phage-inclusive metagenomics profiling was conducted with Phanta.

### Profiling of bacterial structural variations

Prior to the structural variations (SVs) classification, the iterative coverage-based read assignment pipeline was used to precisely reassign the ambiguous reads to the most likely reference(19,20). The reference genomes given in the proGenomes database (http://progenomes1.embl.de/) were concatenated, separated into genomic 1-kbp bins, and used for the detection of highly variable genomic variations. The SGV-Finder pipeline(19) was used to identify SVs. For genomic segments with a deletion frequency below 25% across the population, standardized coverage was computed for that structural variant, namely variable SVs (vSVs). If the deletion frequency falls between 25% and 75%, only the presence or absence of the segment was recorded, namely deletion SVs (dSVs). Regions with a deletion frequency of >75% were excluded from further analysis. All bacterial species with SVs calling were found in at least 10% of all samples and were used in the analysis that followed.

### Enterotype classification

The relative abundances of bacterial taxa were first calculated from the shotgun metagenomic sequencing data. Taxonomic profiling was conducted at the genus level, and only genera with a relative abundance greater than 0.1% in at least 50% of the samples were included in the analysis to reduce noise. We conducted Principal Coordinate Analysis (PCoA) on the Bray-Curtis dissimilarity matrix derived from the genus-level taxonomic profiles. To classify the samples into distinct enterotypes, we applied the Partitioning Around Medoids (PAM) clustering algorithm to the PCoA results. The optimal number of clusters was determined by calculating the silhouette coefficient for different cluster numbers, with the highest silhouette score indicating the best clustering solution. Based on the clustering results, samples were assigned to one of the enterotypes. The dominant genera in each enterotype were identified and used to characterize the enterotype profiles, with specific genera such as Bacteroides and Prevotella serving as key indicators for the respective enterotypes.

### Molecular docking simulation

The crystal structures of several human receptors were obtained from the RCSB Protein Data Bank (https://www.rcsb.org/). These include the free fatty acid receptor 1 (GPR40, PDB ID: 5TZY), the thromboxane A2 receptor (TXA2, PDB ID: 6IIV), and the P2Y purinoceptors 1 and 12 (PDB IDs: 4XNW and 7PP1, respectively). Subsequent to download, water molecules and co-crystallized ligands were removed from the .pdb files to prepare the proteins for molecular docking studies. For the docking simulations, the receptor structures were prepared by adding polar hydrogen atoms and computing partial charges using the Kollman and Gasteiger methods. These modifications were performed using AutoDock Tools (version 1.5.638). The prepared structures of both ligands and receptors were then saved in .pdbqt format, facilitating their use in subsequent molecular docking procedures. The molecular docking studies were performed using the open-source software AutoDock Vina (version 1.1.2) (21) investigate the interactions between AMI-associated long-chain fatty acids (LCFAs) and selected receptors involved in platelet aggregation.

Adjustments to the grid box allowed the AutoDock algorithm to predict ligand conformations within the receptor binding sites and evaluate them using scoring functions (22), with the grid spacing set at 1 Å. The three-dimensional interactions between the selected receptors and long-chain fatty acids (LCFAs) were analyzed and visualized using the PyMOL Molecular Graphics System (version 2.5.0), an open-source platform. Subsequently, AutoDock version 4.2 was employed to predict the binding affinities between various ligands and the selected receptors, providing quantitative insights into the potential binding interactions (23).

### Thrombogenic effect of LCFAs on isolated human platelets

Fresh platelets were obtained from normal human volunteers without any antiplatelet or anticoagulant drugs being used within 2 weeks. Whole blood was collected from the peripheral vein of healthy volunteers into 0.1 vol of ACD buffer (75 mM sodium citrate, 39 mM citric acid, and 135 mM dextrose, pH 6.5). Diluted whole blood was centrifuged at 1500g for 20 minutes at 25 °C, and platelet-rich plasma (PRP) was collected into a fresh tube. Then we diluted PRP in ACD buffer, and centrifuged it at 800g for 10 minutes at 25℃. The platelet pellet was then resuspended in Tyrode’s buffer (137 mM NaCl, 2 mM KCl, 0.34 mM Na_2_HPO_4_, 12 mM NaHCO_3_, 1 mM MgCl_2_, 5.5 mM glucose, 5 mM HEPES, and 0.35% BSA, pH 6.5) at 10^9^ platelets/mL, and kept at 23 °C for up to 4 h before use.

Incubations of washed human platelets with fatty acids were done either in pH 6.5 phosphate buffer or in “pH 7.4 Tris buffer” (15.4 mM Tris-Cl, pH 7.4; 140 mM NaCl, 5.6 mM glucose(24)). Commercially available LCFAs, pre-selected to represent each class of LCFAs, were diluted to specific concentration (1 mM, 2.5mM, 5mM and 10mM) by bovine serum albumin solution (BSA), respectively. Platelet aggregations were measured using a lumi-aggregometer (AG800, Shandong, China) at 37 °C under stirring at 1200 rpm.

### Statistical analysis

In this study, no data were excluded before the statistical analysis. As the study is observational, no allocation or randomization was used. This study includes all available samples (nU=U56) of patients with AMI and healthy individuals. Although sample sizes were not predetermined using statistical methods, our sample sizes are comparable to those published earlier(25,26). The distribution of samples among metagenomics and metabolomics batches was random. During the data collecting phases of the metagenomic, biochemical, and metabolomics, researchers were blinded to the group allocation.

All statistical analyses were performed with R (version 4.2.3). Significantly differed metabolites or taxa between AMI and control groups were found by parametric (student’s *t*-test) or non-parametric (Wilcox’s rank test) statistical methods. Multivariate analysis approaches including principal component analysis (PCA), principal coordinate analysis (PCoA), and random forest (RF) were also conducted. Permutational multivariate ANOVA (PERMANOVA) was performed to determine the differences in structures of global microbiome and LCFAs between AMI patients and controls by using *vegan* (v2.6-4) package. The association analysis between omics features were performed by spearman correlation using *cor.test* base command. Least absolute shrinkage and selection operator (LASSO) regression model was built by using *glmnet* (v4.1-7) package. Unless otherwise stated, all p values were corrected using the Benjamini-Hochberg method andz PadjU<U0.05 was considered statistically significant.

## Results

### Plasma metabolome and gut microbiome from a deeply phenotyped eAMI cohort

We profiled plasma samples collected from 30 early acute myocardial infarction (eAMI) patients (within 12 hours in emergency) and 26 age- and gender-matched healthy controls (HC), using mass spectrometry. For these participants, we also characterized gut microbiota by performing shotgun metagenomic sequencing of fecal samples. In addition, as shown in **Table 1**, it is important to note that HC in this study had no obstructions in in any of their coronary arteries, which was validated by coronary angiography, in addition to having bioclinical characteristics that were within heathy thresholds. As expected, eAMI patients are distinct from HC with elevated troponin, pro-BNP, and creatine kinase (p values < 1.0e-03) despite matching for age, sex, BMI and T2D status. Also, eAMI patients display increased inflammatory markers including C-reactive protein, white blood cells, neutrophils, and lymphocytes (p values < 1.0e-03).

**Table 1.**
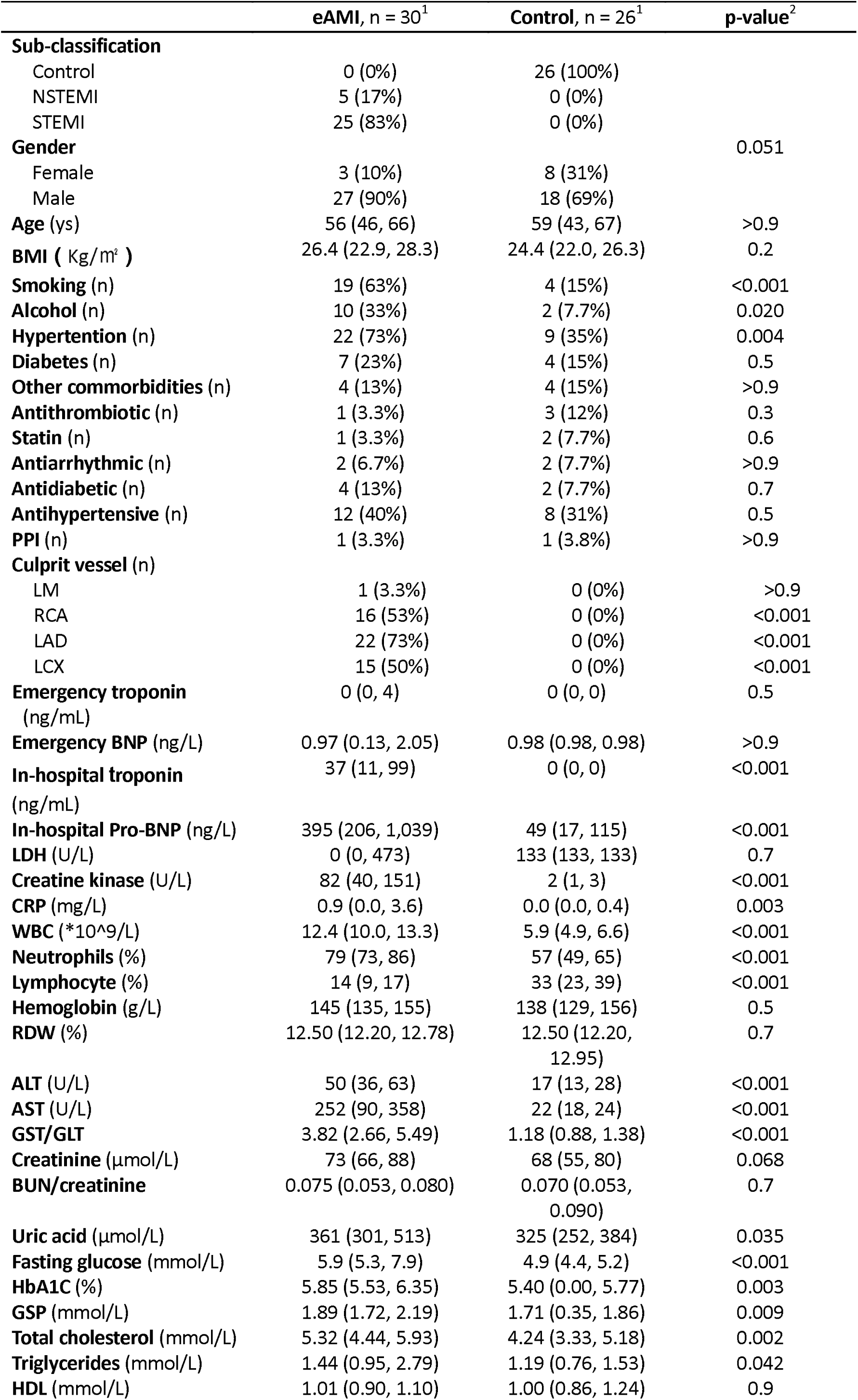
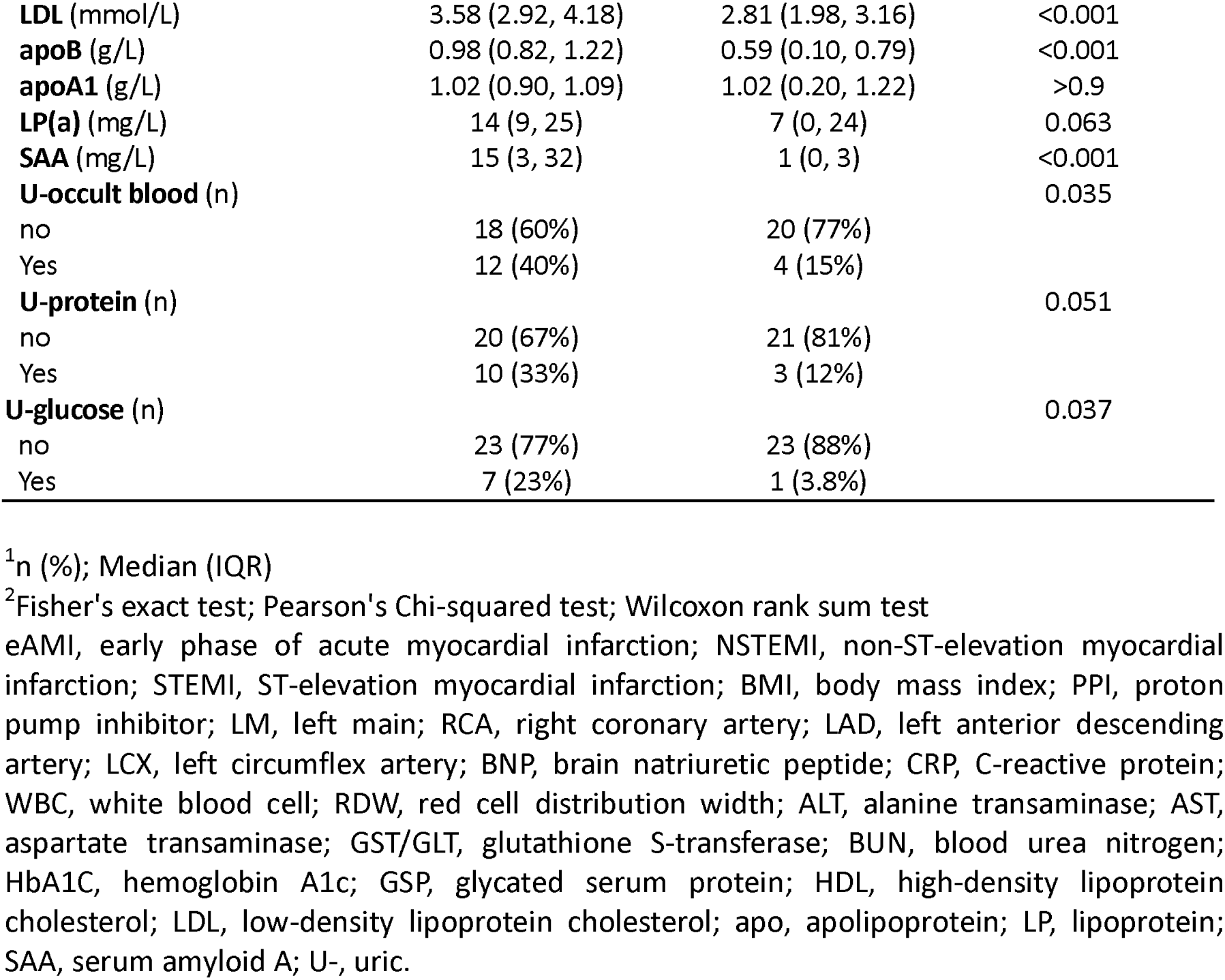
Cohort characteristics.

A pseudo-targeted metabolomics platform was used to profile 926 identified metabolites, as well as 26 partially characterized features (**Table S1**). Metabolites belonged to diverse biochemical classes, including lipids, amino acids, xenobiotics, nucleotides, and carbohydrates (**Figure S2a**). Most metabolites (877) were measured in over 50% of the cohort, and 555 metabolites were present in all samples (**Figure S2b**). All the samples were randomly injected and median of RSDs for the internal standards used in the quality control samples is 2.78%, demonstrating the high quality of metabolomics data.

Bacterial DNA was extracted from fecal samples, sequenced on the Illumina platform, and a total of 402 Gb 150-bp paired-end reads were generated, with an average of 47.70 ± 5.98 (s.d.) million reads per sample (**Table S2**). For each sample, a majority of high-quality sequencing reads (69.03–97.24%) were *de novo* assembled into long contigs or scaffolds, which were used for gene prediction, taxonomic classification, and profiling of structural variations (SVs) in bacterial genomes.

### LCFAs dominate the metabolome perturbations in eAMI plasma

Principal component analysis of the 952 measured compounds is used in **Figure 1a** to show the remarkable differences in the global plasma metabolome of eAMI patients compared to HC subjects and individual contrasted metabolites are shown in **Figure S3**. To characterize the dysregulated metabolic pathways in patients with eAMI, we performed KEGG pathway enrichment analysis of the differential metabolites and observed that biosynthesis of unsaturated fatty acids (FA) was the most significantly disrupted in eAMI patients (**Figure 1b**). Moreover, we found that across the eAMI and HC groups, the 32 LCFAs species (saturated, monounsaturated, and polyunsaturated) showed considerable inter-individual heterogeneity (**Figure 1c**). Plasma LCFAs composition also displayed a significant variation between the two groups (permutational multivariate analysis of variance (PERMANOVA), p < 1.0e-03; **Figure 1d**), with 28 upregulated LCFAs between patients with eAMI compared with HC as demonstrated in the volcano plot (**Figure 1e**).

**Figure 1.**
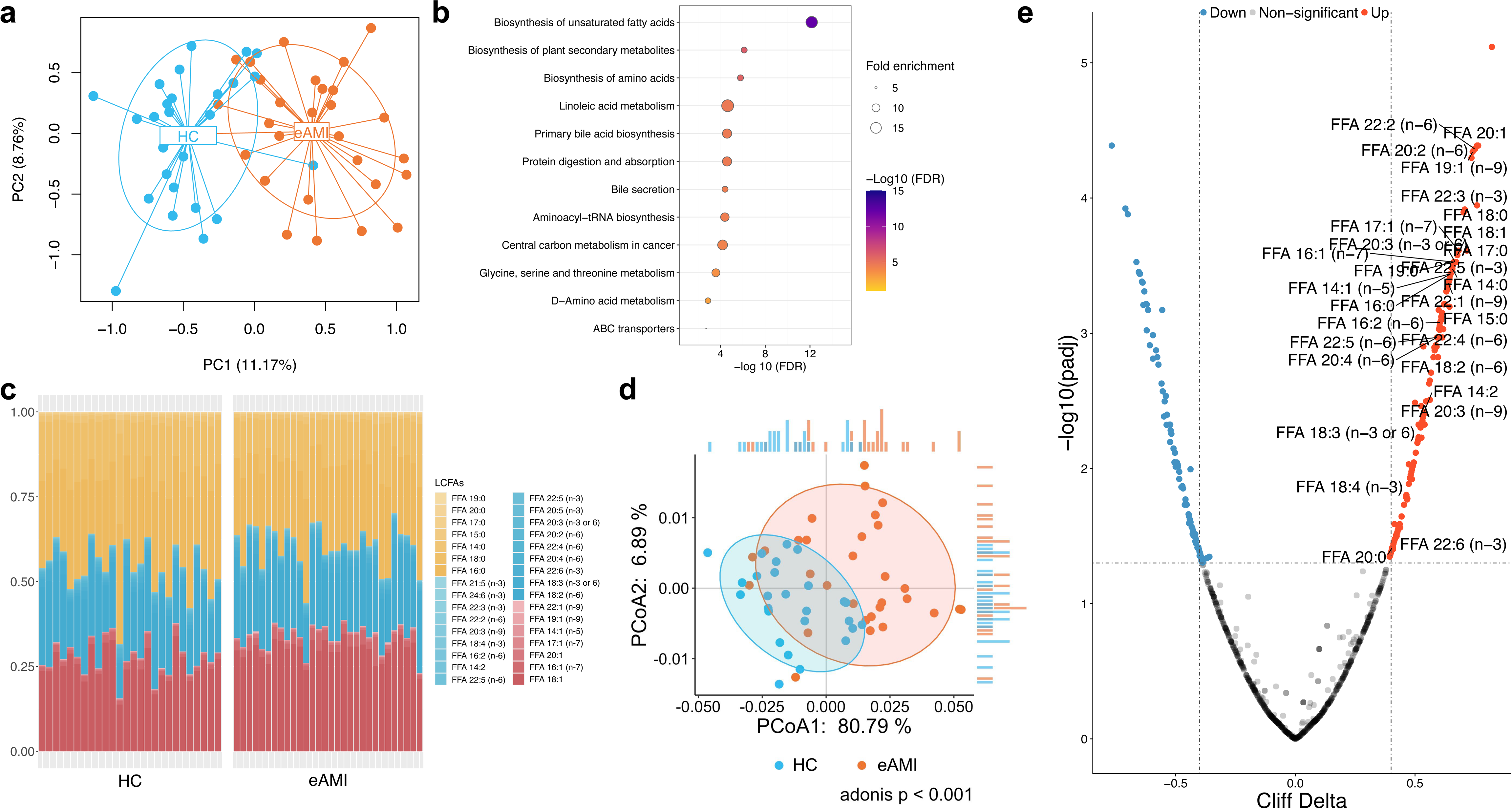
Long-chain free fatty acids (LCFAs) dominate the metabolome alterations in plasma of early acute myocardial infarction (eAMI) patients compared with healthy control (HC) individuals. **(a)** Principal component (PC) analysis demonstrates the remarkable metabolome alterations in eAMI patients compared with HC individuals. **(b)** KEGG metabolic pathway enrichment of differential metabolites (FDR-adjusted p < 0.05) between eAMI and healthy controls. Fisher’s exact test (one-side) followed by FDR-adjusted p value was used and only pathways with FDR-adjusted p < 0.05 were shown. **(c)** Proportional structure of 32 detected LCFAs in plasma across all samples of eAMI and HC individuals. LCFAs are colored by degree of saturation, saturated LCFAs are colored in yellow, monounsaturated LCFAs are colored in red, and polyunsaturated LCFAs are colored in blue. **(d)** Principal coordinate analysis (PCoA) plot of the difference between eAMI and HC groups based on LCFAs proportion profile. **(e)** Volcano plot of 952 detected metabolites by untargeted metabolome profiling in eAMI patients and HC individuals. Red dots represent significantly up-regulated (FDR-adjusted p < 0.05 and Cliff Delta > 0.4) metabolites in eAMI plasma. Blue dots represent notably eAMI-depleted plasma metabolites (FDR-adjusted p < 0.05 and Cliff Delta < −0.4). Grey dots marked the unchanged metabolites. Significantly altered LCFAs were labeled. The horizontal line represents FDR cutoff of 0.05 and the vertical lines denote |Cliff Delta| > 0.4.

Thereafter, we divided our cohort into a training group (comprising 70% of the subjects) and a test set (comprising 30% of the subjects) to identify diagnostic biomarkers for differentiating HC and patients with eAMI. The training dataset was utilized to perform feature selection using LCFAs in plasma, and the selected features were then used to create a random forests classifier (**Figure 2a**). With an area under the receiver operating characteristic curve (AUC) of 0.93 in the training set, the random forests model by a biomarker panel of 9 LCFAs (**Figure 2b**), including FFA 16:0, FFA 22:2 (n-6), FFA 15:0, FFA 24:6 (n-3), FFA 22:6 (n-3), FFA 14:2, FFA 20:3 (n-3 or 6), FFA 20:5 (n-3), and FFA 22:5 (n-6), allowed differentiation between HC and eAMI groups (**Figure 2c**). The created classifier was subsequently used in testing dataset to estimate independent performance of our model, which had an AUC of 0.89 through 10-fold cross-validation (**Figure 2d**). Overall, these findings reveal the differences of LCFAs alteration is dominant in metabolome perturbations of patients with eAMI and demonstrate LCFAs may allow to discover more potent biomarkers, making the diagnosis of eAMI more accessible.

**Figure 2.**
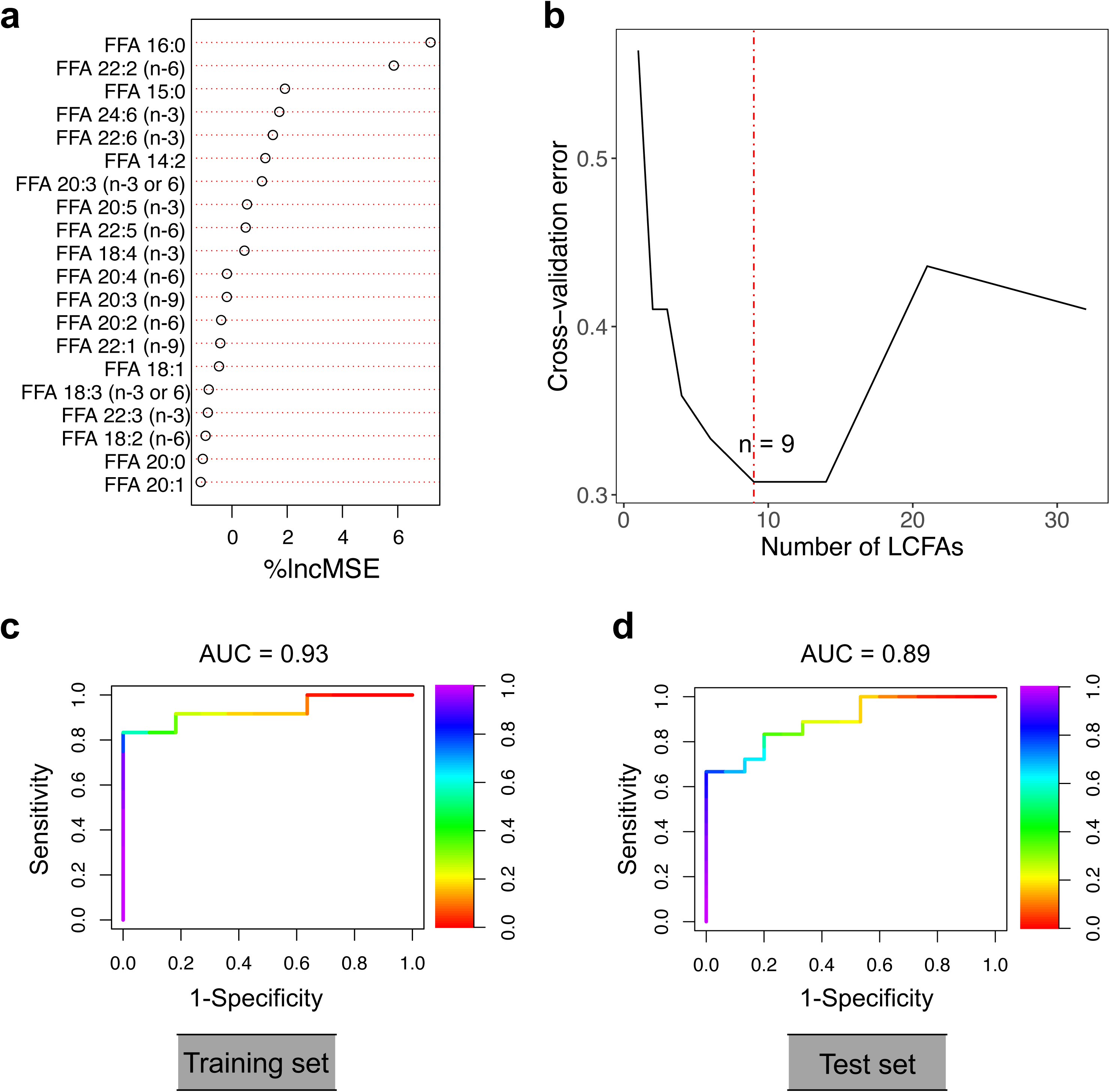
Machine learning demonstrates long-chain free fatty acids (LCFAs) are potential diagnostic biomarkers for eAMI. **(a)** The top 20 most important LCFAs reducing the variance (mean squared error (%IncMSE)) in classification of eAMI by random forest model. %IncMSE denotes the increase in mean squared error of predictions as a result of variable (LCFAs) being permuted. The training model was constructed by proportional profile of 32 LCFAs in 39 training samples. **(b)** Distribution of cross-validation error in random forest classification of eAMI patients as the number of top ranking LCFAs increased. Red dot line marks the optimal number of LCFAs (*n* = 9) for the classification of eAMI patients. **(c and d)** Receiver operating characteristic (ROC) curve using the 14 LCFAs optimized by random forest model for the (**c**) training and (**d**) test set (right panel, *n* = 9). Vertical heatmap in each panel denotes specificity for the ROC curve in c and d. AUC, area under the curve.

### Alterations in gut microbiome is associated with the LCFA dysmetabolism in eAMI

We next explored the underlying factors that regulate LCFA metabolism by factor analysis, where we found age, sex, and body mass index could only account for up to 10% of the variance in LCFA proportion (**Figure 3a**). This indicates that a large proportion of LCFA variation remains unexplained and may be attributed to other variables, for example, gut microbial factors. Consistent to previous reports(12), we identified that eAMI gut microbiome is not only distinct from but also more heterogenous than that of HC subjects (**Figure S4**). Furthermore, we found eAMI gut microbiome is marked by an enrichment in the Bacterides2 enterotype (*P*_Fisher’s exact test_U=U2.3e-02) with a slight depletion in Bacteroides1 enterotype (*P*_Fisher’s exact test_U=U3.1e-01, **Figure 3b**), which always represents a marker suggesting a high risk of metabolic disorders(27). In addition, we observed a minor decrease in the *Ruminococcus* enterotype in the eAMI group (*P*_Fisher’s exact test_ = 5.7e-03).

**Figure 3.**
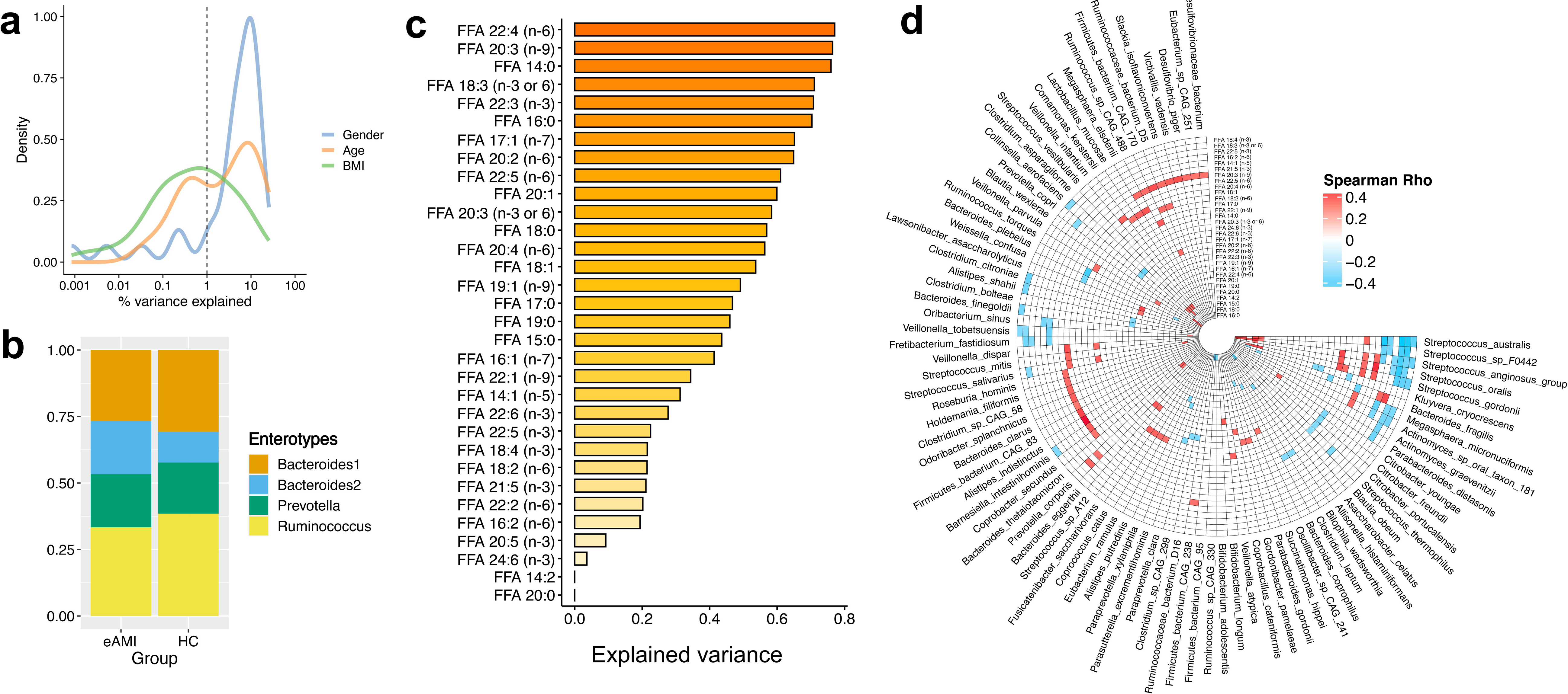
Bacterial alterations characterize eAMI gut microbiome and greatly explain the variance in plasma long-chain free fatty acids (LCFAs) . **(a)** Density plot showing sample-wide distribution of (% of explained variances) intrinsic factors (gender, age, and body mass index (BMI)) associated with the plasma concentrations of LCFAs. **(b)** Bar plot demonstrates the variation of 32 LCFAs explained by identified bacterial taxa at species level using LASSO regression. **(c)** Stacked bar plot shows the prevalence of gut enterotypes in eAMI and control groups. **(d)** Circular heatmap represents the correlation between LCFAs and gut bacterial species. Only association groups with FDR-adjusted p value of < 0.01 are colored in this plot.

To furtherly understand if gut microbiome is involved in the regulation of LCFAs during the onset of eAMI, we performed a least absolute shrinkage and selection operator (LASSO) regression model to compute the variance in each of the detected LCFAs that can be explained by gut microbiome. Intriguingly, gut microbiome at species level explains up to 70% of the variance in LCFAs composition in this cohort (**Figure 3c**). We next sought to identify the potential bacterial taxa that associate with the LCFAs by spearman correlation. At genus level, *Mycobacteroides*, *Thermus*, and *Anaerostipes* are inversely linked to LCFAs (**Figure S5**). As shown in **Figure 3d**, we found *Streptococcus* species were negatively correlated with omega-3 polyunsaturated FAs including FFA 18:4 (n-3), FFA 18:3 (n-3 or −6), and FFA 22:5 (n-3), which were considered as cardioprotective compounds. Meanwhile, the *Streptococcus* species were found to be positively correlated with omega-6 polyunsaturated FAs and saturated LCFAs. Moreover, we discovered that the eAMI gut virome exhibits a decrease in *Vibrio* phage (**Figure 4a**) but enrichments in *Streptococcus* phage PH10 and *Streptococcus* phage SM1 (**Figure 4b**), both of which have known hosts that are *Streptococcus* species. *Streptococcus* bacterial species predominate in the eAMI gut microbiome (**Figure S4a**), although the mechanism underpinning this dominance is unclear. Nonetheless, the abundance in temperate lysogenic *Streptococcus* phages may provide some justification. Additionally, the total number of virulent phages found in the eAMI gut microbiome is higher than in the healthy controls (**Figure 4c**). The ratio of virulent to temperate phages is also greater in the eAMI group compared to the HC group.

**Figure 4.**
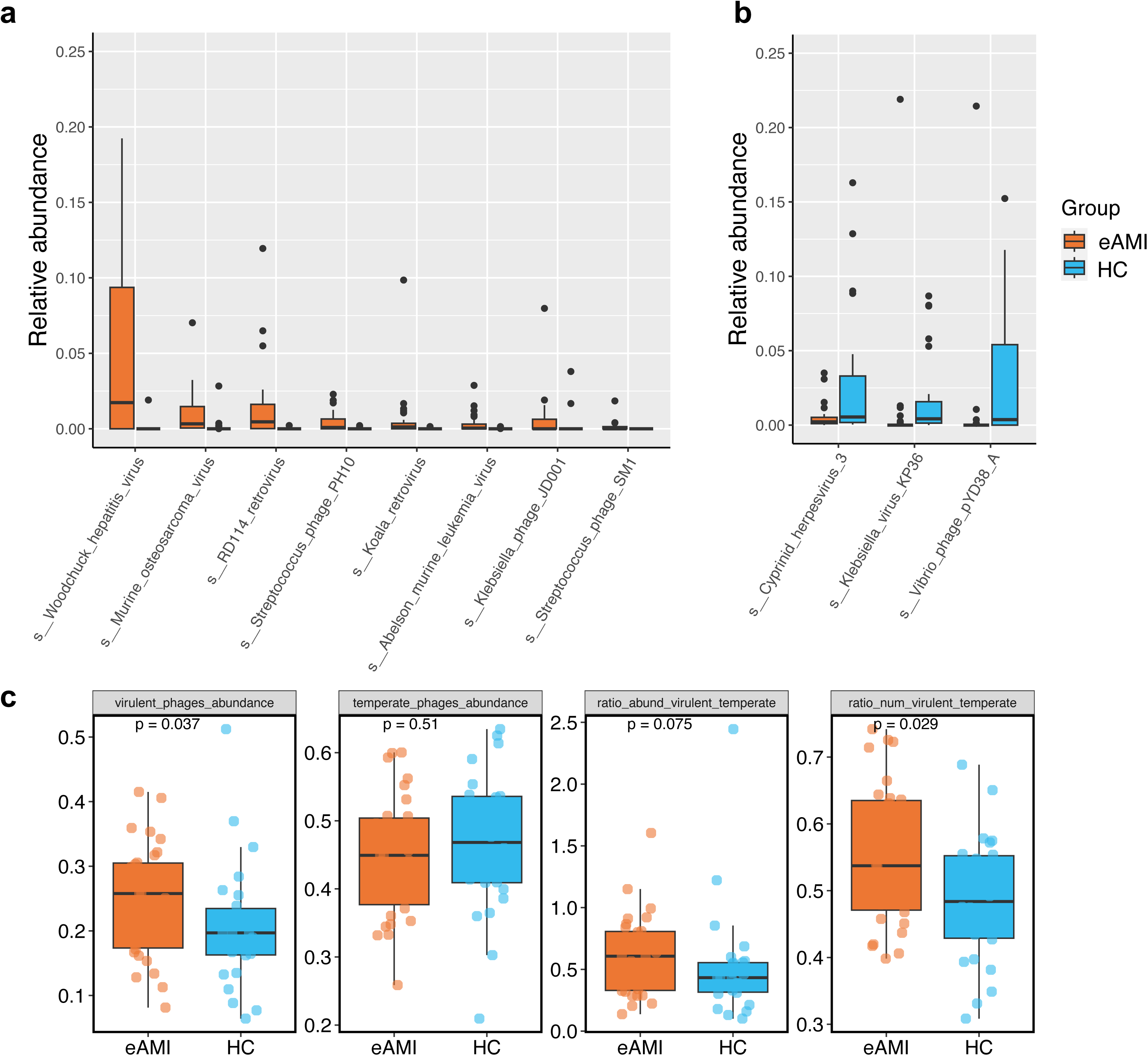
Alterations in viral gut microbiota between eAMI and HC groups. Box plots showing the relative abundance of increased **(a)** and decreased **(b)** viral gut microbiota at species level in eAMI gut microbiome compared to that of HC group. Differences in abundance were detected using MaAslin2. **(c)** Significant variations in the abundance, abundance ratios, and counts of virulent and temperate phages between the HC and eAMI groups. Significance was determined by Wilcoxon ranked sum test. For box plots in all panels, the vertical lines extend 1.5 times the interquartile range (top and bottom borders of the box) and the median depicted by the horizontal line inside the box.

### Gut bacterial structural variants profiling in eAMI

Structural variations (SVs) in bacterial genomes can disrupt gene function, thereby influencing the interactions between bacteria and their hosts (25). As a result, significant phenotypic and functional alterations within the gut microbiome of patients with early acute myocardial infarction (eAMI) may arise from differences in the presence or levels of SVs among genetically similar bacterial strains. Here, we profiled SVs in all samples and identified 4,234 deletion SVs and 1,948 variable SVs across 47 bacterial species (**Figures 5a** and **5b**). For some species, we observed considerable differences in copy number variation. We identified 87 deletion SVs and 18 variable SVs in *Bacteroides uniformis* across 134 individuals, 78 deletions and 15 variable SVs in *Faecalibacterium prausnitzii* in 124 individuals, and 110 deletion SVs and 55 variable SVs in *Parabacteroides distasonis* in 121 individuals. For the archaeal microbiota, we only identified SVs in *Methanobrevibacter smithii* in 13 individuals (**Figures 5a**). To explore potential differences in bacterial genetics, we further computed the Canberra distance of bacterial SV profiles between eAMI versus HC (**Figure 5c**). eAMI and HC samples were significantly different in the beta-diversity of SV composition (*P*_Wilcoxon_ = 1.4e-05) and composition of bacterial genetics in eAMI individuals has less heterogeneity than that of HC subjects.

**Figure 5.**
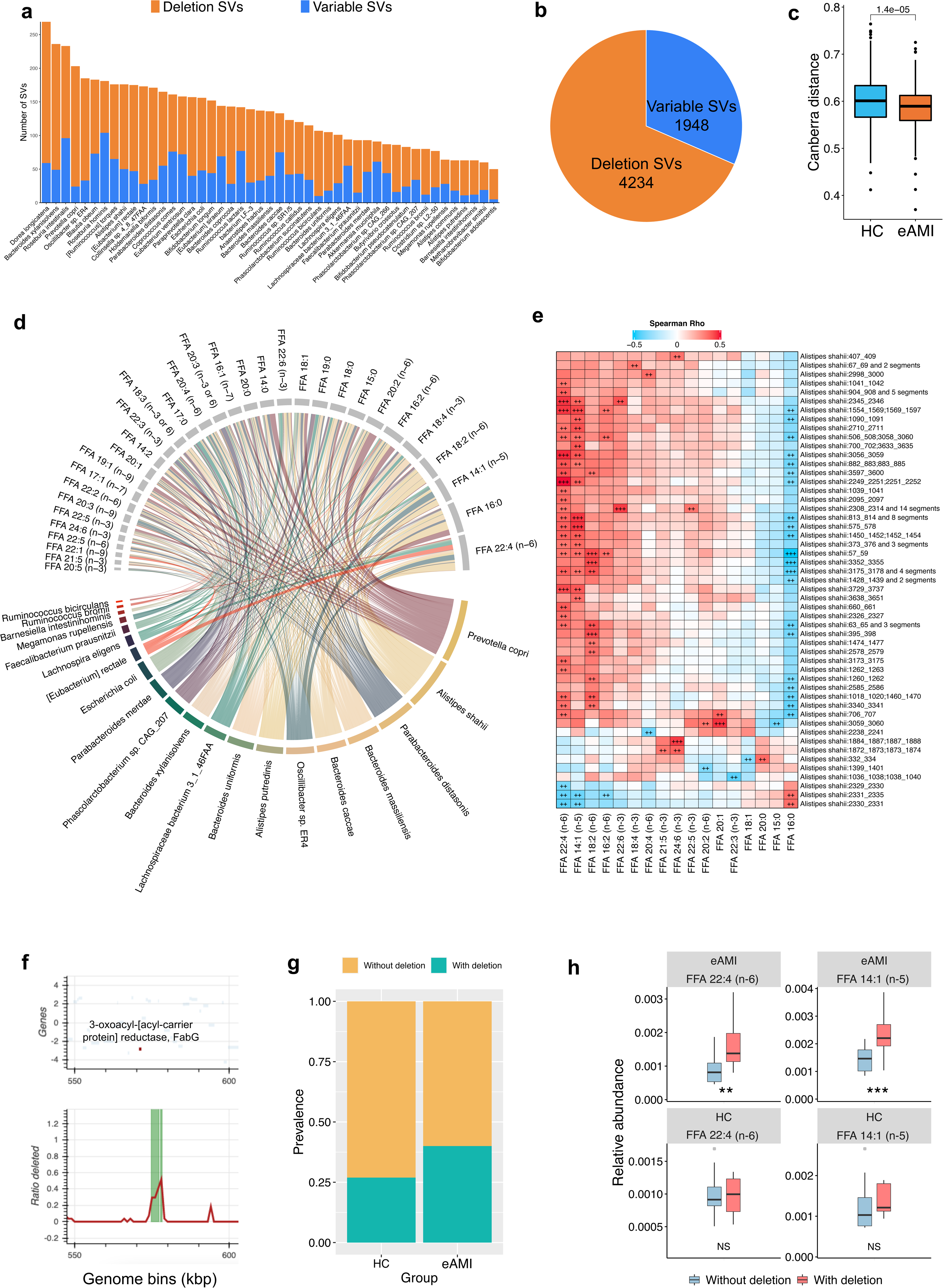
Bacterial structural variations (SVs) links to long-chain free fatty acids profiles in acute myocardial infarction (AMI) patients. **(a)** Number of SVs of each bacterial species in 56 study participants. **(b)** Pie chart showing the total identified SVs numbers. **(c)** Boxplot of Canberra distance between samples within eAMI and healthy control (HC) groups based on SVs profile, significance is determined by two-sided Mann-Whitney *U*-test. **(d)** Chord diagram showing significant associations between LCFAs and bacterial SVs. **(e)** Heatmap demonstrates the associations between SVs of *Alistipes shahii* and LCFAs. Only significant correlations with FDR-adjusted p value of < 0.01 are visualized. ++, FDR-adjusted p < 0.01, and +++, FDR-adjusted p < 0.001. **(f)** The deletion rate of the 3-kbp deletion harboring 3-oxoacyl-[acyl-carrier protein] reductase (FabG) in *Alistipes shahii*. **(g)** Stacking bar plot represents the prevalence of 3-kbp deletion SV of *Alistipes shahii* in eAMI and HC groups. **(h)** Relative abundance of LCFAs in individuals with and without the 3-kbp deletion SV of *Alistipes shahii* in eAMI (upper panel) and HC (bottom panel) groups. **, p < 0.01, ***, p < 0.001 determined by two-sided Mann-Whitney *U*-test.

### Bacterial genetics associate to LCFAs dysmetabolism in eAMI

Exploring relationships between gut bacterial SVs and plasma LCFAs, we found that bacterial SVs were significantly related to LCFAs (**Figure 5d**). As a particularly notable example, in the association analysis between SVs in the *Alistipes shahii* genome and plasma LCFAs profile, we found that a 3-kbp deletion (*A. shahii*:575-578) was directly associated with FFA 22:4 and FFA 14:1 but inversely linked to FFA 16:0, indicating that this deletion SV may be involved in the regulation of LCFAs metabolism in eAMI (**Figure 5e**). According to gene analysis, the beginning of this specific genomic region in *A. shahii* encodes a 3-oxoacyl-[acyl-carrier protein] reductase (*FabG*, **Figure 5f**), which is an enzyme catalyzes the NADPH-dependent reduction of beta-ketoacyl-ACP substrates to beta-hydroxyacyl-ACP products, the first reductive step in the elongation cycle of fatty acid biosynthesis. This deletion region shows a trend toward higher prevalence in eAMI group compared to HC gut microbiome (**Figure 5g**). In accordance with the association analysis, we furtherly found that eAMI individuals with *A. shahii* genomes harboring this deletion have higher plasma FFA 22:4 and FFA 14:1, compared to eAMI participants without this 3-kbp deletion in A. *shahii* genome (**Figure 5h**). Interestingly, no such differences were observed for any of the FFAs in the HC group, suggesting a specific interaction between the genomic deletion and FFA metabolism in eAMI participants.

### Molecular docking and thrombosis in human platelets support the thrombogenic effect of eAMI-associated LCFAs

We extended our work to *in silico* molecular docking to investigate the molecular connections that control thrombosis. Four canonical receptors that are involved in platelet aggregation—free fatty acid receptor 1, also known as G-protein coupled receptor 40 (GPR40), thromboxane A2 receptor (TXA2), P2Y purinoceptor 1 (P2Y1), and P2Y purinoceptor 12 (P2Y12)—were taken into account as binding receptors in the molecular docking models in addition to the 28 eAMI-associated LCFAs as binding ligands. As shown in **Figure 6a**, we selected the first-ranked complex models between the various LCFAs and these four receptors. Interestingly, the molecular docking models estimated substantial interactions between eAMI-associated LCFAs and four selected receptors, with binding affinity ranging from −5.7 to −9.1 kJ/mol (**Figure 6a**). Representative docking results for bindings between the chosen receptors and omega-3, −6, and −9 polyunsaturated FAs are displayed in **Figure 6b**.

**Figure 6.**
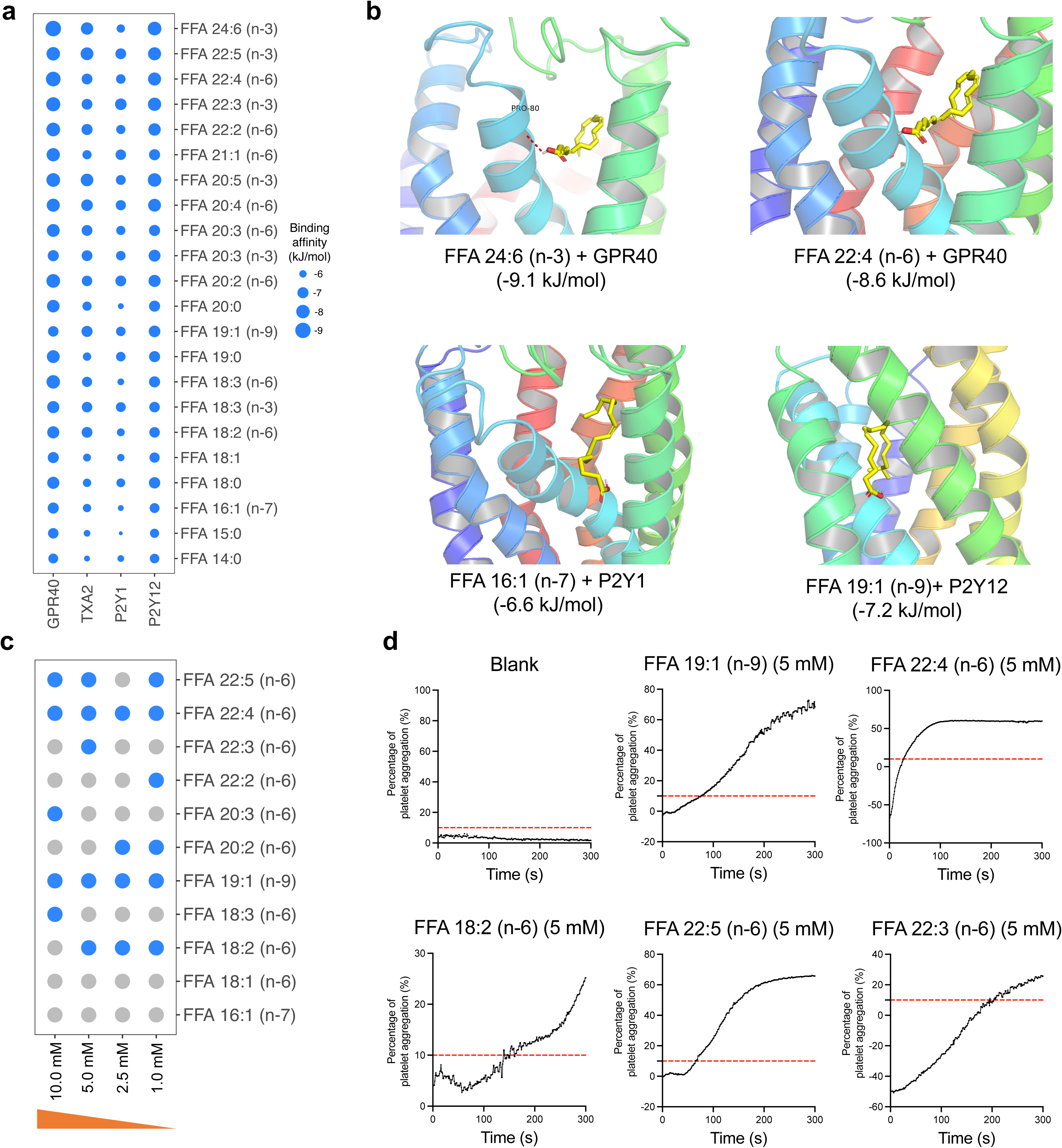
Early acute myocardial infarction (eAMI)-associated long-chain free fatty acids (LCFAs) bind to thrombosis-related receptors and induce thrombosis in human platelets at physiological relevant concentrations. **(a)** Heatmap of binding energies between eAMI-associated LCFAs and thrombosis-related receptors through molecular docking. Bing affinities are quantified at kJ/mol. **(b)** The representative AutoDock predicted binding conformation complex of LCFAs versus thrombosis-related receptors, within which the receptors are depicted as cartoons and LCFAs as sticks. **(c)** The thrombogenic impact of eAMI-associated LCFAs on human platelets is depicted in a heatmap. Platelet aggregation ratios of > 10% and 10% are indicated by blue and grey, respectively. **(d)** Representative platelet aggregation curves in a time interval of 300 seconds after the treatment of LCFAs at a concentration of 5 mM. Red dashed line indicates 10% threshold.

Then, using *in vitro* human platelet anticoagulation assay, we aimed to experimentally validate the thrombogenic effects of eAMI-associated LCFAs. As shown in **Figure 6c**, we found omega-6 LCFAs significantly stimulate platelet aggregation at physiological concentrations (1 mM to 10 mM), reflecting their overall reactivity rather than a dose-dependent effect. As a non-ignorable example, FFA 19:1 (n-9), as a non-omega-6 LCFA, also causes strong thrombosis in human platelet (**Figure 6d**), while the mechanistic understanding behind this finding remains unknown. Our results imply suggest that eAMI-associated LCFAs may possess potent thrombogenic functionality, making it fair to assume that they play a significant role in host cardiometabolic health.

## Discussion

The factors triggering the onset of AMI and the precise indicators of eAMI remain unclear. However, the application of plasma metabolomics provides valuable insights into the search for accurate diagnostic markers for complex conditions such as AMI. Additionally, comprehensive multi-omics profiling plays a crucial role in unraveling the underlying mechanisms responsible for AMI. Together, these approaches shed light on both diagnostic advancements and a deeper dissection disentangling the complexity of eAMI.

By plasma metabolomics profiling, we found that in patients with eAMI, there was a remarkable increase in 28 LCFAs dominating the altered eAMI metabolome. These LCFAs encompassed a variety of saturated, monounsaturated, and polyunsaturated types. When investigating the etiology of LCFA dysmetabolism in eAMI, we initially examined its correlation with conventional AMI risk factors including age, sex, body mass index, and potentially diabetes. However, these factors alone could only account for up to 10% of the observed variation in LCFA proportions. This observation led us to explore other potential contributors, focusing on the gut microbiota due to its recognized role in metabolome regulation. Using a LASSO regression model, we found that the gut microbiome at the species level accounted for up to 70% of the observed variance in LCFA composition. This finding highlights a strong association between the gut microbiome and the changes in LCFAs levels, although it does not establish a causal relationship.

Our findings indicated a notable shift in the gut virome composition in eAMI patients, particularly with enrichments in Streptococcus phage PH10 and Streptococcus phage SM1. These phages were known to target Streptococcus species, which we found to be positively correlated with omega-6 polyunsaturated FAs and saturated LCFAs. This association suggested that the alterations in the gut virome may influence the abundance of Streptococcus species, potentially contributing to the dysregulation of FAs metabolism observed in eAMI. While the exact mechanisms remain to be fully elucidated, these findings underscore the complex interplay between the gut microbiota, virome, and metabolome in eAMI, offering new insights into how microbial dynamics may influence disease progression.

To gain mechanistic insights behind the gut microbiome-LCFA interactions, we associated bacterial SVs with LCFAs and observed significant relations. Of specific interest is a 3-kbp deletion (*A. shahii* : 575-578) representing *FabG* gene, a key enzyme in reduction step of elongation cycle in fatty acid biosynthesis, which is more prevalent in eAMI gut microbiome, resulting in a relatively higher plasma FFA 22:4 and FFA 14:1 in eAMI patients. Others have already reported the increase of intracolonic saturated LCFAs positively associated with elevated abundances of *Prevotella, Lactobacillus*, and *Alistipes*(28). Additionally, the presence of *A. shahii* was also shown to strengthen the positive associations of red meat intake with trimethylamine N-oxide (TMAO) concentrations levels(29), while increased TMAO levels enhances platelet hyperreactivity and thrombosis risk(30), and is associated with an increased risk of incident major adverse cardiovascular events(31). These findings support that in addition to short-chain fatty acids – which are known gut microbial products – disturbances in the gut microbiota also contribute significantly to the variation in plasma LCFAs during eAMI.

Indeed, the question arises whether the elevated levels of LCFA in eAMI are merely a marker or if they hold pathophysiological significance. Considering that platelet activation is a distinct pathophysiological characteristic of eAMI, we sought to explore the impact of eAMI-associated LCFA on circulating platelet activation. As expected, we found a majority of omega-6 LCFAs rather than omega-3 LCFAs stimulate platelet aggregation. A role for omega-3 polyunsaturated fatty acids (PUFA) supplementation to achieve cardioprotective effects has also been shown by attenuating platelet function(32), aligning with our findings. To date, there is limited conclusive evidence regarding the effect of LCFAs on platelet activation, except for arachidonic acid (FFA 20:4 (n-6)), a well-known LCFA that activates platelets. However, our study has yielded intriguing results. We have discovered that both FFA 19:1 (n-9) (also known as 10-nonadecenoic acid, HMDB0013622) and FFA 22:4 (n-6) (Adrenic acid, HMDB0002226) can significantly induce human platelet aggregation to a similar degree as arachidonic acid (FFA 20:4 (n-6), see **Figure S6**). Notably, FFA 22:4 (n-6) can be synthesized from arachidonic acid (FFA 20:4 (n-6)), which, in turn, can be elongated from FFA 18:2 (n-6), also known as linoleic acid. These conversions and interactions among omega-6 PUFAs provide a potential mechanism for their role in activating platelets.

Consistent with previous reports(33), the outcome of our *in vitro* experiments supports many eAMI-associated LCFAs, especially omega-6 PUFAs can promote platelet aggregation even in normal physiological concentration (1-5 mM). However, lots of epidemiological investigation has found higher intakes of omega-6 PUFA appear to be safe and may be even more beneficial, hence the AHA supports an omega-6 PUFA intake of at least 5% to 10% of energy for dietary recommendations of coronary heart disease(34). In another prospective analyses, higher *in vivo* circulating and tissue levels of LA and possibly AA were associated with lower risk of major cardiovascular events(35). Intriguingly, omega-6 PUFA, dihomo-γ-linolenic acid (FFA 20:3 (n-6)) has been shown to play a role in inhibiting platelet aggregation ex vivo, via COX-1–derived prostanoid metabolites(36). Therefore, it still has the overwhelming controversial data on potential effects of omega-6 PUFAs on platelet function.

## Limitations of the study

While our study offered valuable multi-omics insights into a cohort of patients with eAMI, uncovering the association between gut dysbiosis and plasma metabolomic perturbations characterized by the notable enrichment of LCFAs, it is important to acknowledge certain limitations in our present study. Firstly, while our study offers important insights into the relationship between gut dysbiosis and fatty acid dysmetabolism in eAMI, it is primarily correlative. Further studies, including longitudinal and interventional designs, are needed to confirm the causal pathways suggested by our findings. Secondly, while platelet activation is a characteristic feature of eAMI, the specific *in vivo* role of LCFAs is still not well understood, and there is ongoing debate regarding the effect of ω-6 LCFAs on platelet function. Thirdly, we still don’t know the direct or indirect effect of gut microbiota dysbiosis on platelet activation, as well the underlying mechanism responsible for it, despite there is report on TMAO(30) and phenylacetylglutamine(37) contributing to platelet hyperreactivity and enhanced thrombosis potential. Fourthly, although all participants followed a standardized in-hospital diet and stool samples were collected shortly after hospitalization, the potential influence of pre-hospital diet on intestinal microbiome and plasma metabolome profiling, as well as their interaction, cannot be completely ruled out. Finally, due to the limitations of sample size and the specific population background included in this study, the conclusions drawn from our findings should be furtherly validated through large-scale, prospective studies involving diverse regional populations.

## Data availability

The metadata are available under restricted access due to participant consent and privacy regulations of this cohort, access can be obtained by request to the corresponding author (Hanbin Cui). Shotgun metagenomic sequencing reads of the gut microbiome is available at National Genomics Data Center of China *via* accession ID PRJCA018032 and metabolomic raw LC-MS data has been deposited at MetaboLights with accession ID MTBLS11092.

## Code availability

Scripts associated with the data analysis and visualization are available at https://github.com/fjw536/AMI_gut_microbiome.

## Acknowledgements

We thank the study participants and nurses for their contributions to the project. We gratefully acknowledge colleagues at Lianchuan biotechnology for Metagenomics sequencing analysis of gut microbiota, and DI’AN Diagnostics, Key Laboratory of Digital Technology in Medical Diagnostics of Zhejiang Province for plasma metabolomics profilling, data analysis and discussions.

## Author contributions

Y.F., J.Y. and H.C. conceived and designed the study. J.Y., F.H., K.H., T.H., P.S. X.H., H.H., W.L., J.L., J.C., S.W., R.Z., H.M., F.H., J.S., H.Z., Z.L., X.F., X.S., P.W., M.G., W.D., S.L., Y.W., F.Y., R.F., Z.K., and X.C. were involved in the acquisition of samples and data. Y.F. and Z.K. analyzed and interpreted the data. Y.F. and J.Y. drafted the work and J.Y. and H.C. substantively revised it. All authors contributed to the article and approved the submitted version.

## Supplementary Figures

**Figure S1.**
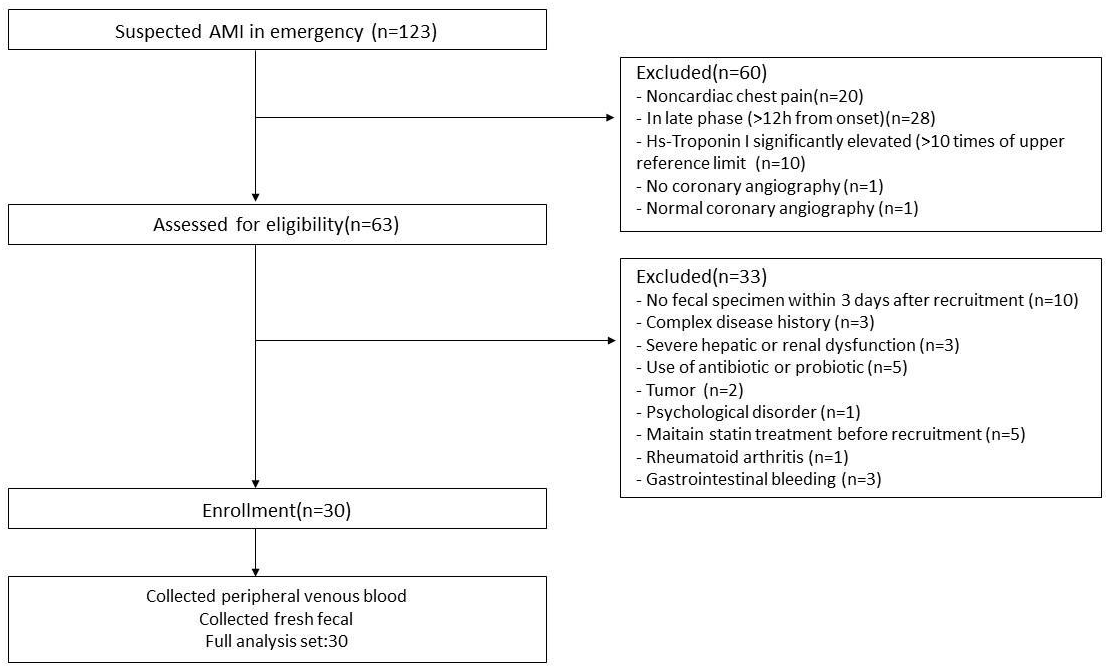
Participant flow diagram. 123 suspected myocardial infarction patients were contacted, and 30 were eventually enrolled after screening.

**Figure S2.**
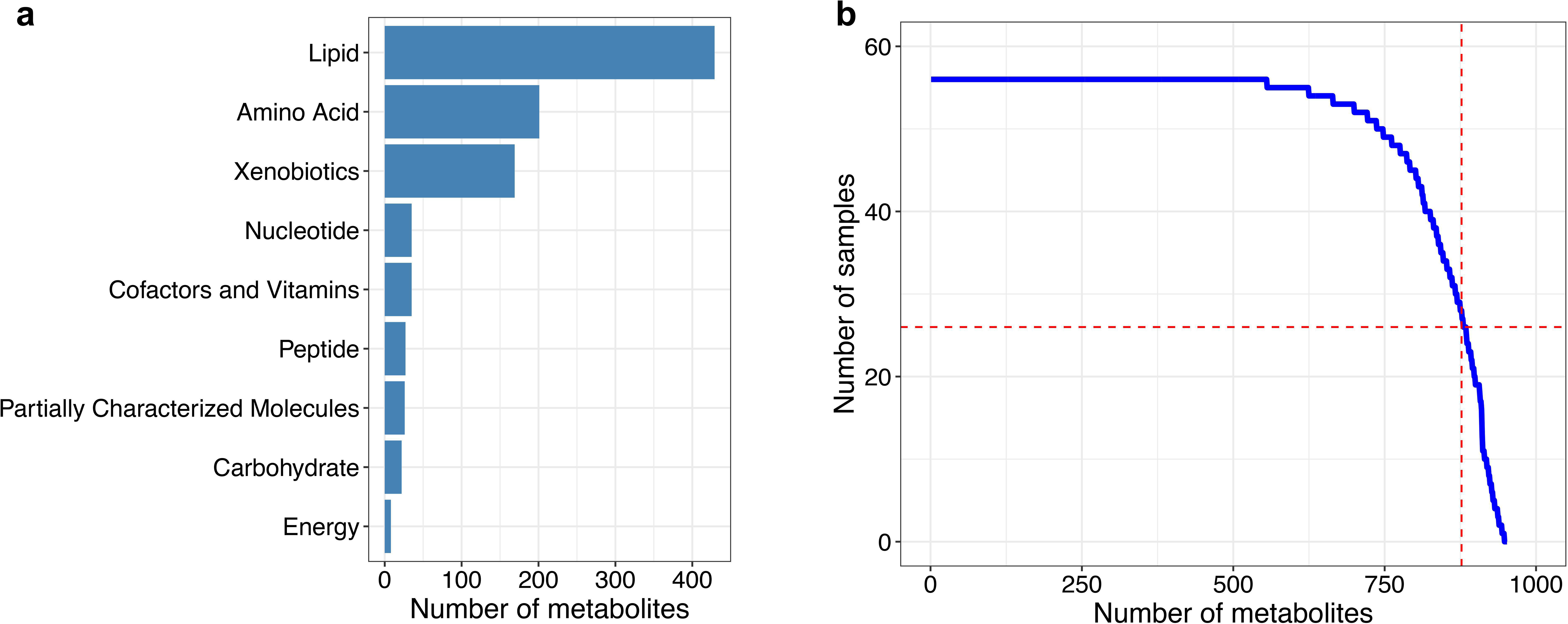
**a**, Distribution of measured metabolite super pathways of the 952 metabolites. **b**, Distribution of metabolite prevalence among the 56 samples. Blue distribution represents prevalence of all analyzed metabolites (*n*U=U952). Numbers of metabolites that are present in more than 50% (*n* = 28) of the samples are indicated by dashed lines.

**Figure S3.**
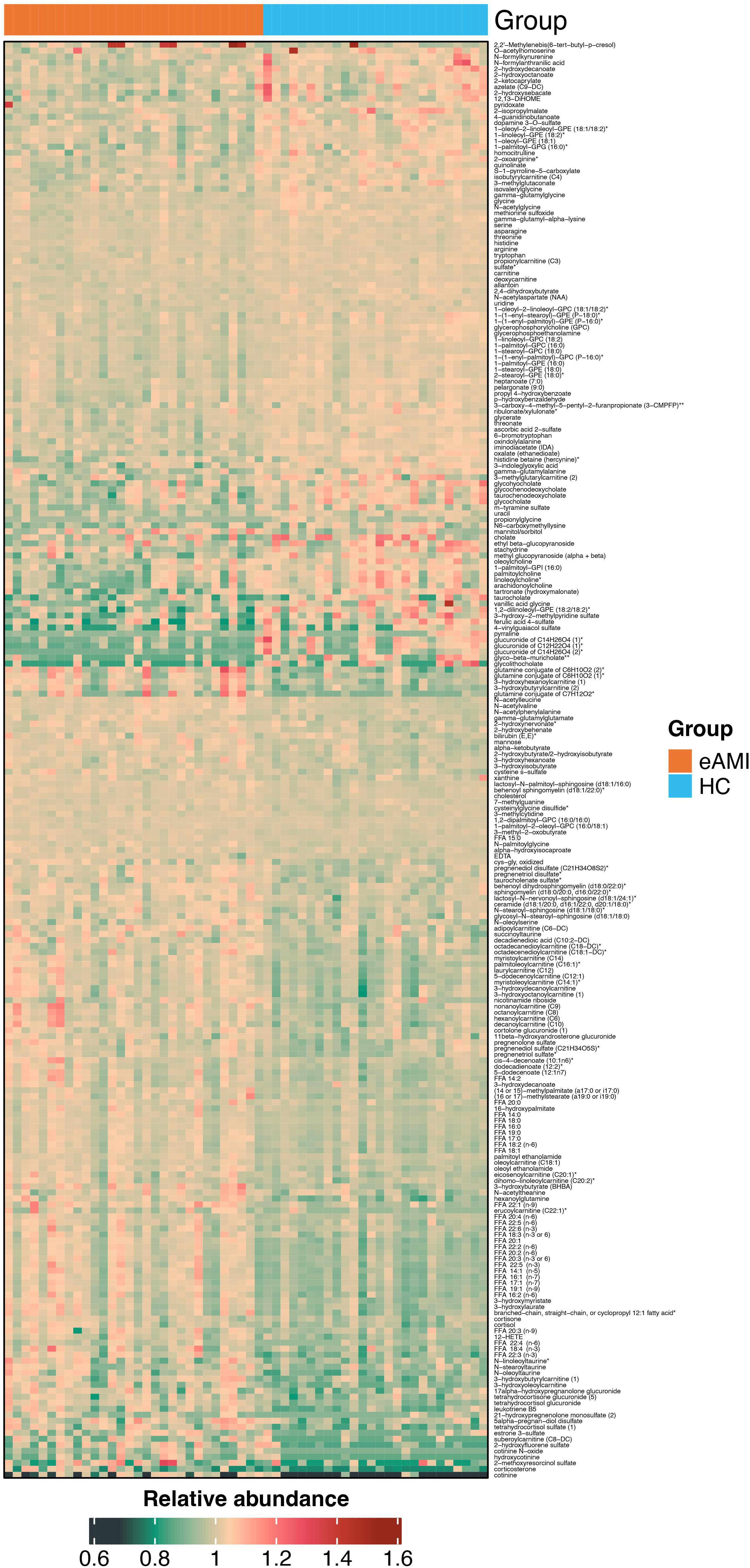
Metabolic alterations in the plasma of eAMI patients. Heatmap of contrasted metabolites in the plasma of healthy controls (HC) and eAMI patients.

**Figure S4.**
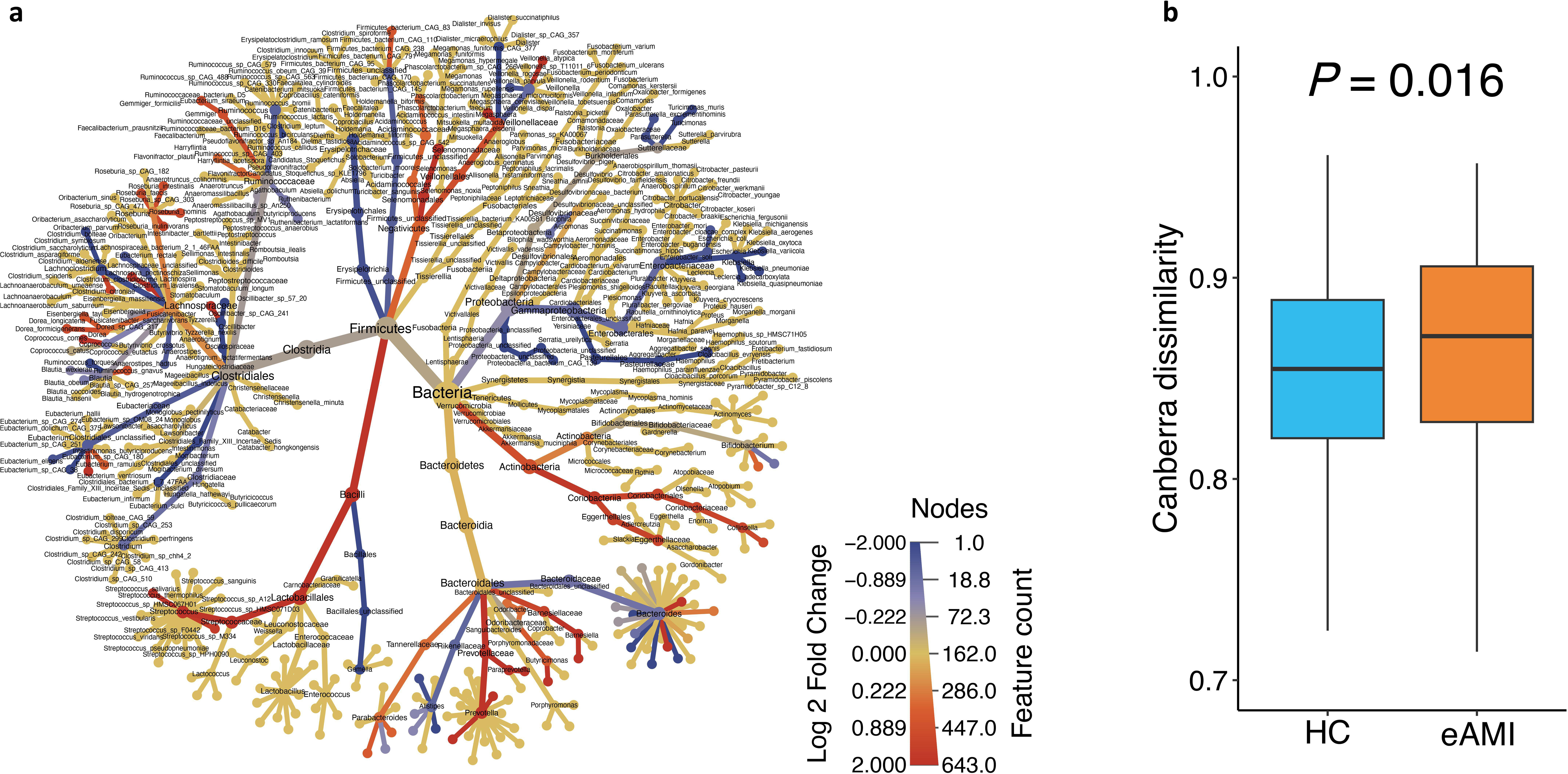
Alterations in bacterial gut microbiota in eAMI patients. **a**, Phylogenetic tree of contrasted bacterial taxa between eAMI and HC bacterial gut microbiota. Taxa colored in red and blue respectively represents increased and reduced taxa in eAMI bacterial gut microbiota compared with HC group. **b**. Box plot (line, median; box, interquartile range (IQR); whiskers, 1.5× IQR) of β-diversity at bacterial species level (Canberra distance) of eAMI (*n* = 30) and HC (*n* = 26) gut microbiota. Statistical significances between two groups were determined by Wilcoxon rank-sum test (two-sided).

**Figure S5.**
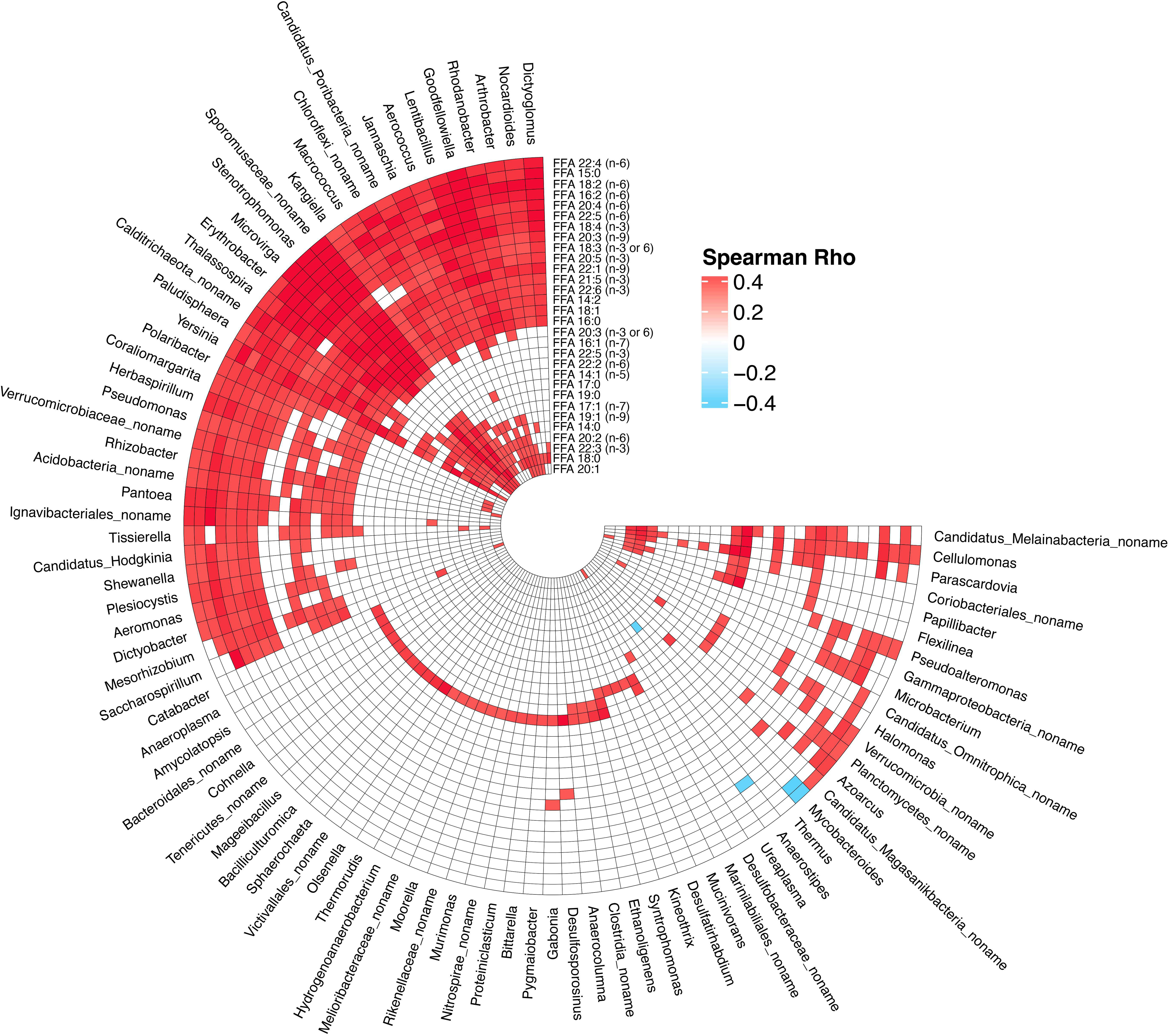
Gut microbiome at genus level associate with long-chain fatty acids (LCFAs) metabolism. Circular heatmap represents the correlation between LCFAs and gut bacteria at genus level. Association groups with FDR-adjusted p value of < 0.05 are visualized in this plot.

**Figure S6.**
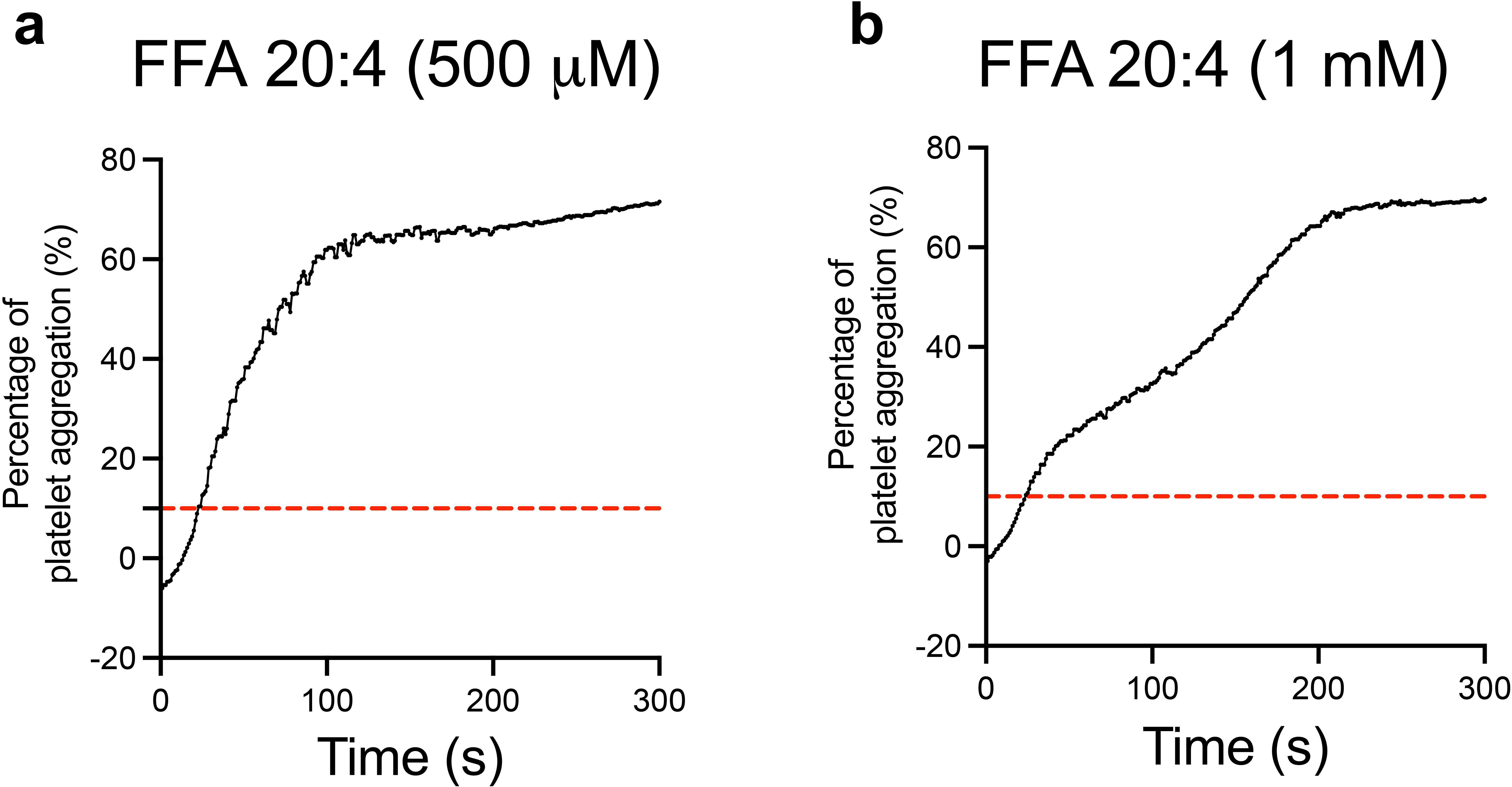
Thrombogenic effects of FFA 20:4 (arachidonic acid) in human platelets. Platelet aggregation curves in a time interval of 300 seconds after the treatment of FFA 20:4 at concentrations of 500 µM and 1 mM are given in **(a)** and **(b)**, respectively. Red dashed line indicates 10% threshold.

